# Predicting mortality in critically ill patients with hypertension using machine learning and deep learning models

**DOI:** 10.1101/2024.08.21.24312399

**Authors:** Ziyang Zhang, Jiancheng Ye

## Abstract

**Background:** Accurate prediction of mortality in critically ill patients with hypertension admitted to the Intensive Care Unit (ICU) is essential for guiding clinical decision-making and improving patient outcomes. Traditional prognostic tools often fall short in capturing the complex interactions between clinical variables in this high-risk population. Recent advances in machine learning (ML) and deep learning (DL) offer the potential for developing more sophisticated and accurate predictive models.

**Objective:** This study aims to evaluate the performance of various ML and DL models in predicting mortality among critically ill patients with hypertension, with a particular focus on identifying key clinical predictors and assessing the comparative effectiveness of these models.

**Methods:** We conducted a retrospective analysis of 30,096 critically ill patients with hypertension admitted to the ICU. Various ML models, including logistic regression, decision trees, and support vector machines, were compared with advanced DL models, including 1D convolutional neural networks (CNNs) and long short-term memory (LSTM) networks. Model performance was evaluated using area under the receiver operating characteristic curve (AUC) and other performance metrics. SHapley Additive exPlanations (SHAP) values were used to interpret model outputs and identify key predictors of mortality.

**Results:** The 1D CNN model with an initial selection of predictors achieved the highest AUC (0.7744), outperforming both traditional ML models and other DL models. Key clinical predictors of mortality identified across models included the APS-III score, age, and length of ICU stay. The SHAP analysis revealed that these predictors had a substantial influence on model predictions, underscoring their importance in assessing mortality risk in this patient population.

**Conclusion:** Deep learning models, particularly the 1D CNN, demonstrated superior predictive accuracy compared to traditional ML models in predicting mortality among critically ill patients with hypertension. The integration of these models into clinical workflows could enhance the early identification of high-risk patients, enabling more targeted interventions and improving patient outcomes. Future research should focus on the prospective validation of these models and the ethical considerations associated with their implementation in clinical practice.

## INTRODUCTION

Hypertension is a prevalent chronic condition that significantly increases the risk of severe complications, including heart disease, stroke, and kidney failure.[1] Critically ill patients with hypertension admitted to the Intensive Care Unit (ICU) often present with complex medical profiles and are at a heightened risk of mortality.[2] Early and accurate prediction of mortality in these patients is crucial for guiding clinical decision-making, optimizing resource allocation, and improving patient outcomes. However, predicting mortality in this population is challenging due to the multifactorial nature of their conditions and the dynamic, rapidly changing clinical environment of the ICU.[3]

Traditional prognostic tools, such as scoring systems like the Acute Physiology and Chronic Health Evaluation (APACHE) and Sequential Organ Failure Assessment (SOFA), have been widely used to assess the severity of illness and predict outcomes in ICU patients.[4] While these tools provide valuable insights, they are often limited by their reliance on a predefined set of clinical variables and may not fully capture the complex interactions between patient characteristics and clinical outcomes. Furthermore, the predictive accuracy of these traditional models can be compromised by the heterogeneous nature of critically ill populations, including those with hypertension.[5]

Recent advances in machine learning (ML) and deep learning (DL) have opened new avenues for developing more sophisticated and accurate predictive models.[6] These models can handle large, multidimensional datasets and automatically identify complex patterns and interactions within the data, offering the potential for more personalized and precise predictions.[7] In particular, deep learning models, such as convolutional neural networks (CNNs) and recurrent neural networks (RNNs), have shown promise in various clinical applications, including the prediction of mortality, disease progression, and treatment response.[8, 9]

Despite the potential advantages of ML and DL models, their application in predicting mortality among critically ill patients with hypertension remains underexplored. This study seeks to address this gap by evaluating the performance of various ML and DL models in predicting mortality in this high-risk population. Specifically, we compare traditional ML models, such as logistic regression and decision trees, with advanced DL models, including 1D CNNs [10] and long short-term memory (LSTM) networks.[11] We also investigate the importance of different clinical features in predicting mortality, with a particular focus on the role of commonly used ICU scoring systems, such as the APS-III score and SOFA score.[12]

The objectives of this study are twofold: first, to identify the most effective predictive model for mortality in critically ill patients with hypertension; and second, to provide insights into the key clinical features that drive these predictions. By leveraging the strengths of both ML and DL approaches, we aim to contribute to the development of more accurate and clinically useful prognostic tools that can support decision-making in the ICU.

## METHODS

### Dataset processing

This study utilized the Medical Information Mart for Intensive Care IV (MIMIC-IV) dataset, which contains de-identified healthcare information for approximately 40,000 patients admitted to critical care units at Beth Israel Deaconess Medical Center (BIDMC) between 2008 and 2019. [13] Patients with hypertension were identified using ICD-9 and ICD-10 codes.[14] We extracted the following data from the patients’ electronic health records (EHRs): (1) Demographics, including self-reported race, sex, and age; (2) Vital signs; (3) Laboratory test results; (4) Maximum creatinine levels on days two and three; and (5) Patient mortality.

We also calculated the Sequential Organ Failure Assessment (SOFA) Score and the Acute Physiology Score III (APS-III) for each patient. The SOFA score assesses organ failure in ICU patients, tracking the status of six organ systems: respiratory, cardiovascular, hepatic, coagulation, renal, and neurological.[15] SOFA is advantageous as it does not require specific tests and relies on routinely collected ICU data. The APS-III score evaluates the severity of illness in adult ICU patients [16] and is an enhanced version of the Acute Physiology and Chronic Health Evaluation (APACHE) system.[17] APS-III is designed to predict hospital mortality with greater accuracy by assessing the patient’s current health status, underlying medical conditions, and complications arising during their ICU stay. It is widely used in critical care research to compare illness severity across patients and to adjust outcomes in clinical studies. [18]

We addressed missing data using Multivariable Imputation by Chained Equations (MICE).[19] MICE is a robust statistical technique that imputes missing values with plausible estimates, creating complete datasets. Initially, missing values are filled with preliminary imputations, such as means and standard deviations. The method then iteratively models each variable with missing data as a function of other variables in a chained equation system, updating the imputations at each step. This process is repeated over multiple cycles until convergence. Multiple complete datasets are generated, analyzed separately, and the results are combined to account for imputation uncertainty.

This study aimed to predict patient mortality using various predictors and to examine the relationship between these predictors and mortality. We employed multiple machine learning models, including Decision Trees and Support Vector Machines (SVM), as well as deep learning models, such as one-dimensional convolutional neural networks (1D CNNs) and recurrent neural networks (RNNs). We excluded data with values outside reasonable ranges (e.g., heart rate above 200).

### Statistical analysis

Statistical analysis was conducted using Python and the open-source library “statsmodels”.[20] The analysis had two main objectives: (1) to explore the bivariate association between patient mortality and predictors, and (2) to perform initial feature selection using conventional statistical machine learning models, specifically multivariable logistic regression. We defined a model with all predictors as the initial selection and a model with highly associated predictors as the backward selection. For the initial selection, we performed multivariable logistic regression to obtain unadjusted relationships with p-values. Predictors with the highest p-values greater than 0.05 were iteratively removed until all remaining variables had p-values below 0.05. The resulting set of predictors was considered strongly relevant and used as input variables for the backward selection. Model performance was assessed using root mean square error (RMSE) through 10-fold cross-validation. For logistic regression, odds ratios, p-values, and 95% confidence intervals were computed. Receiver operating characteristic (ROC) curves were drawn for both the initial and backward selections during the 10 runs of cross-validation, and the area under the curve (AUC) was calculated for each run. A higher AUC (closer to 1) indicates better model performance, while an AUC of 0.5 suggests no discrimination.[21]

### Model

We developed multiple classifiers to predict mortality using scikit-learn [22] for machine learning models and TensorFlow [23] for deep learning models. Categorical variables, such as race, were encoded as vectors using one-hot encoding, and continuous variables, such as lab test results, were standardized by subtracting the mean and dividing by the standard deviation to facilitate model fitting.

Several algorithms were used to predict mortality, including logistic regression, support vector machines, multilayer perceptrons (a type of artificial neural network), random forests, bagging decision trees, and boosting decision trees. For logistic regression, we optimized the binary cross-entropy loss using the “Newton-Cholesky” solver, which is well-suited for binary classification with a large number of samples compared to features. For the support vector machine, we employed the stochastic gradient descent algorithm to minimize the log-loss function. The multilayer perceptron architecture included two hidden layers, with 7 neurons in the first layer and 4 neurons in the second, using Rectified Linear Unit (ReLU) activation functions and the Adam solver.[24] For the random forest, we set the number of estimators to 100 and the maximum number of features per tree to the square root of the total features. The bagging decision trees also had 100 estimators, with decision trees as the base estimator. Lastly, the boosting decision trees were implemented using a histogram-based gradient boosting strategy, which is more powerful and efficient for large datasets than other strategies.

To ensure generalizability and robustness, we employed three prominent deep learning methods: 1D CNNs, RNNs, and long short-term memory networks (LSTMs). The 1D CNN, a variant of the traditional 2D CNN, is specifically designed for processing sequential data, such as time-series data. Our model architecture included two 1D convolutional layers with a max-pooling layer in between, followed by two fully connected layers, a dropout layer for regularization, and a SoftMax output layer for classification. For the RNN and LSTM models,[11] we replaced the 1D CNN with RNN and LSTM units, respectively, while keeping the rest of the architecture the same. The binary cross-entropy loss was optimized using the Adam optimizer with a learning rate of 0.0001 over 20 epochs. To avoid overfitting, we monitored the training process by calculating the loss on the validation set and saved the optimal model at the point where validation loss began to increase. In our study, each sample from the dataset was treated as having 36 time steps, with a single feature per time step. Additionally, we incorporated SHAP (SHapley Additive exPlanations) values into our predictor selection process to evaluate predictor importance and refine the backward selection.[25] SHAP is a game-theoretic approach that explains the output of machine learning models by connecting optimal credit allocation with local explanations using the classic Shapley values from game theory.

We evaluated all models using multiple metrics, including AUC, accuracy, sensitivity, specificity, positive predictive value (PPV), and negative predictive value (NPV). The dataset was split into a training set (80%), validation set (10%), and testing set (10%). Hyperparameter tuning was conducted on the training and validation sets, and model performance was evaluated on the test set.

### Study Approval

This study exclusively used publicly available MIMIC-IV data.

## RESULTS

### Statistics overview

A total of 30,096 patients met the inclusion criteria for this study. The demographic and clinical characteristics of these patients are summarized in **Table 1**. The majority of the cohort was white (67.55%), with 13,619 (45.25%) patients deceased following ICU admission, and 16,477 (54.75%) patients surviving. This near-equitable distribution indicates that the classification task is reasonably balanced.

**Table 1.** Characteristics of patients with hypertension in ICU.

| **Variables** | **Total** | **Survival** | **Mortality** | **P-value** |
| --- | --- | --- | --- | --- |
| Sex, N (%) |  |  |  | 0.229 |
| Male | 16985 (56.44) | 9351 (56.75) | 7634 (56.05) |  |
| Female | 13111 (43.56) | 7126 (43.25) | 5985 (43.95) |  |
| Race, N (%) |  |  |  | <0.001 |
| White | 20329 (67.55) | 11058 (67.11) | 9271 (68.07) |  |
| African American | 2804 (9.32) | 1319 (8.01) | 1485 (10.90) |  |
| Hispanic | 1038 (3.45) | 640 (3.88) | 398 (2.92) |  |
| Other | 5925 (19.68) | 3460 (21.00) | 2465 (18.10) |  |
| Age, years, median (IQR) | 67 [57, 78] | 65 [54, 75] | 71 [61, 80] | <0.001 |
| Maximum creatinine during day 2 and day 3, mg/dL | 1.69 (1.69) | 1.45 (1.53) | 1.99 (1.83) | <0.001 |
| Minimum creatinine, mg/dL | 1.47 (1.50) | 1.26 (1.36) | 1.72 (1.61) | <0.001 |
| Maximum creatinine, mg/dL | 1.69 (1.76) | 1.46 (1.62) | 1.97 (1.88) | <0.001 |
| Maximum heart rate, bpm | 128.19 (12.20) | 127.43 (12.15) | 129.12 (12.20) | <0.001 |
| Mean heart rate, bpm | 86.70 (13.84) | 85.06 (13.64) | 87.49 (14.03) | <0.001 |
| Minimum systolic bp, mmHg | 93.46 (18.97) | 96.14 (12.77) | 90.50 (18.66) | <0.001 |
| Mean systolic bp, mmHg | 117.07 (18.45) | 118.66 (18.50) | 115.32 (18.24) | <0.001 |
| Minimum diastolic bp, mmHg | 47.76 (12.83) | 50.01 (12.77) | 45.62 (12.42) | <0.001 |
| Mean diastolic bp, mmHg | 64.14 (14.63) | 65.75 (14.51) | 62.36 (14.55) | <0.001 |
| Minimum SpO2, % | 85.52 (3.29) | 85.54 (3.47) | 85.49 (3.04) | 0.215 |
| Mean SpO2, % | 86.06 (1.76) | 86.07 (1.84) | 86.05 (1.65) | 0.262 |
| Minimum hemoglobin, g/dL | 9.83 (2.24) | 10.08 (2.29) | 9.53 (2.13) | <0.001 |
| Minimum prothrombin, g/dL | 15.70 (7.16) | 14.62 (5.71) | 17.01 (8.41) | <0.001 |
| Maximum prothrombin, g/dL | 17.78 (11.06) | 16.50 (9.12) | 19.34 (12.86) | <0.001 |
| Minimum respiratory rate, bpm | 11.90 (4.68) | 11.55 (4.61) | 12.32 (4.73) | <0.001 |
| Maximum respiratory rate, bpm | 28.91 (8.27) | 28.46 (8.64) | 29.45 (7.75) | <0.001 |
| Mean respiratory rate, bpm | 19.58 (4.07) | 19.15 (3.87) | 20.11 (4.24) | <0.001 |
| Maximum glucose, mg/dL | 165.24 (97.99) | 160.58 (8.98) | 170.85 (106.80) | <0.001 |
| Minimum platelet count, K/μL | 195.95 (109.42) | 195.46 (103.99) | 196.53 (115.67) | 0.408 |
| Minimum calcium, mg/dL | 8.16 (0.85) | 8.17 (0.82) | 8.14 (0.88) | 0.001 |
| Minimum bicarbonate, mg/dL | 22.88 (4.20) | 22.77 (3.82) | 23.02 (4.64) | <0.001 |
| Maximum potassium, mg/dL | 4.49 (0.77) | 4.45 (0.74) | 4.52 (0.80) | <0.001 |
| Maximum blood urea nitrogen, mg/dL | 30.78 (23.56) | 25.45 (19.67) | 37.25 (26.13) | <0.001 |
| Maximum red blood cell count, x 10<sup>6</sup>/μL | 3.63 (0.70) | 3.75 (0.69) | 3.50 (0.70) | <0.001 |
| Minimum red blood cell count, x 10<sup>6</sup>/μL | 3.33 (0.76) | 3.14 (0.76) | 3.24 (0.74) | <0.001 |
| Mean red blood cell count, x 10<sup>6</sup>/μL | 3.49 (0.70) | 3.58 (0.69) | 3.37 (0.70) | <0.001 |
| Maximum white blood cell count, K/μL | 14.09 (10.87) | 13.92 (8.55) | 14.30 (13.14) | 0.004 |
| Maximum sodium, mmol/L | 139.80 (5.87) | 139.80 (5.30) | 139.80 (6.49) | 0.989 |
| Minimum sodium, mmol/L | 128.57 (27.85) | 130.46 (2.43) | 126.24 (31.54) | <0.001 |
| Mean sodium, mmol/L | 135.93 (9.78) | 136.47 (8.50) | 135.28 (11.12) | <0.001 |
| Length of ICU stay, days | 6.00 (6.20) | 5.45 (5.45) | 6.67 (6.93) | <0.001 |
| SOFA score | 4.86 (3.35) | 4.23 (3.08) | 5.62 (3.50) | <0.001 |
| APS-III score | 46.31 (19.64) | 40.74 (17.14) | 53.06 (20.35) | <0.001 |

**Supplemental Table 1** presents the bivariate associations between patient mortality and the predictors of interest. In the unadjusted logistic regression analysis, variables such as minimum SpO2, mean SpO2, minimum platelet count, and maximum sodium levels (p > 0.05) were not significantly associated with patient mortality post-ICU admission. The multivariable logistic regression model using backward selection is detailed in **Supplemental Table 2**. The analysis indicates that patients with elevated red blood cell counts and creatinine levels are more likely to succumb upon ICU admission.

### Models

We conducted logistic regression analyses using 10-fold cross-validation across 10 iterations, each with different random seeds. The results, shown in **Supplemental Table 3**, suggest that the initial selection model exhibits superior classification performance compared to the backward selection model. This finding implies that incorporating all predictors may lead to more accurate predictions.

**Table 2** outlines the performance metrics on the testing set across the models employed in the backward selection, where the top fifteen predictors ranked by the SHAP framework were used as input. Each metric provides unique and critical insights into the model’s performance. Among the models, the 1D CNN achieved the highest AUC (0.7744), demonstrating both good sensitivity and a strong positive predictive value. This suggests that the model not only excels at identifying positive cases but also produces highly reliable positive predictions. These results indicate that the models effectively fit the data without overfitting, due to the cross-validation strategy employed. Notably, the LSTM model reached the highest NPV (0.7317). Overall, the 1D CNN model from the initial selection outperformed the other models across all metrics, indicating that deep learning approaches may be particularly powerful for this study.

**Table 2.**
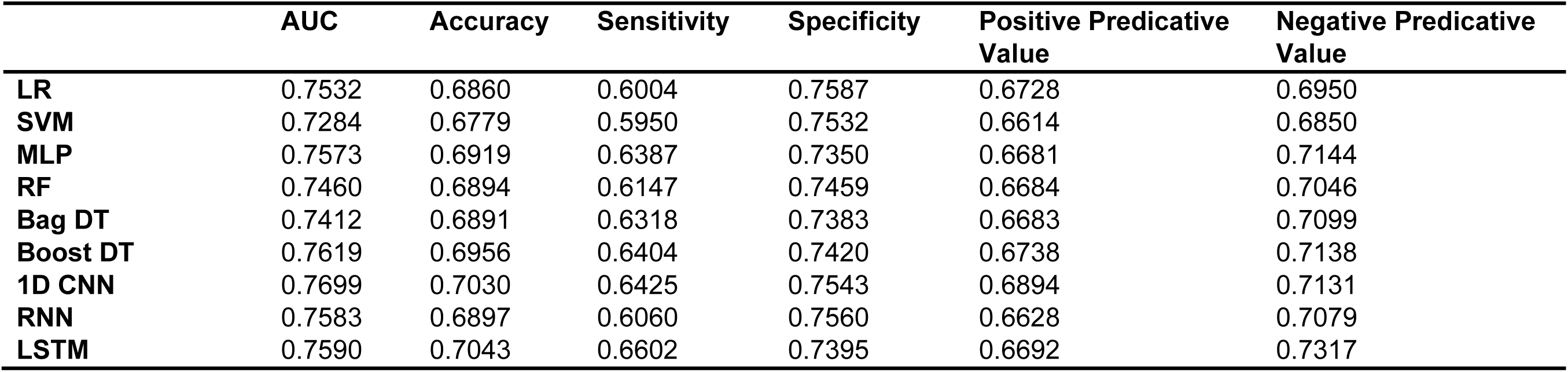
Performance metrics on the models.

Figure 1 illustrates the ROC curves for the machine learning models. Among these, the boosting decision trees model achieved the best AUC (0.762), regardless of whether the initial or backward selection was used. Figure 2 presents the ROC curves for the deep learning models, with the 1D CNN emerging as the top performer (AUC: 0.770).

**Figure 1.**
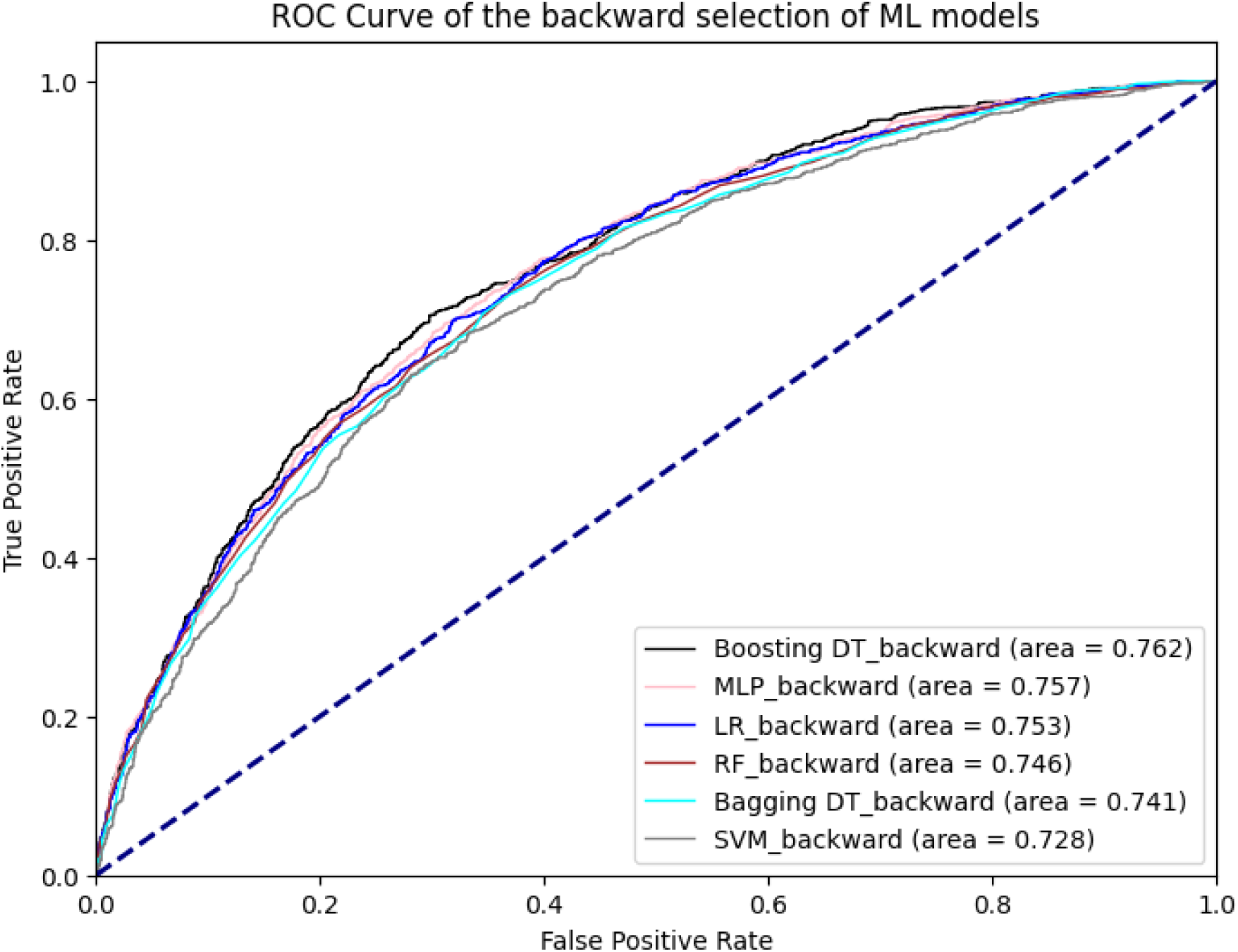
ROC curve of machine learning models.

**Figure 2.**
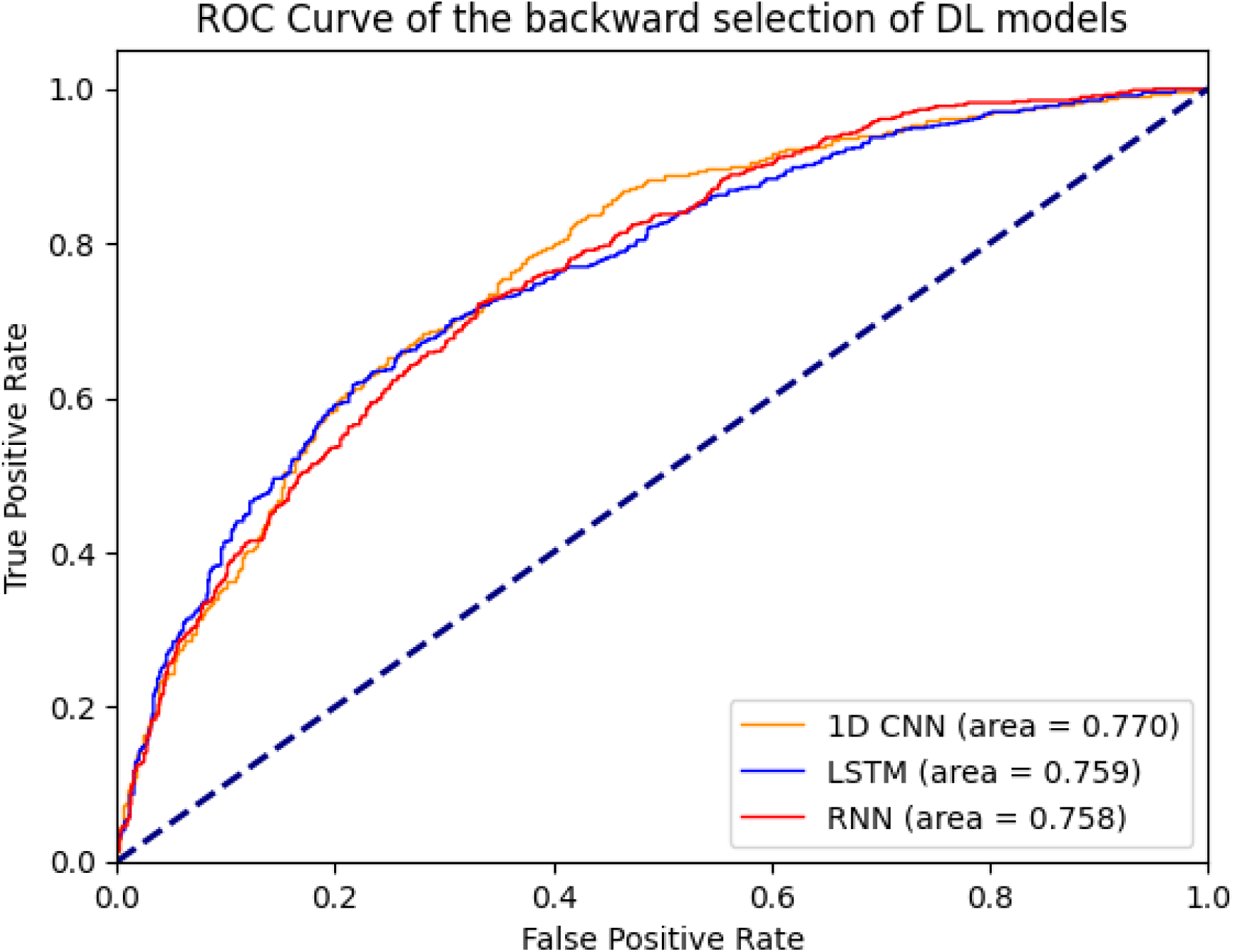
ROC curves of Deep learning models.

### Model interpretation

We provided an interpretation of the 1D CNN model, which had the best overall performance (AUC: 0.774). First, we identified the top 25 predictors based on their mean absolute SHAP values. Figure 3 shows the relationship between these predictors and their influence on the model’s output. Globally, the APS-III score, age, and length of ICU stay were the top three predictors.

**Figure 3.**
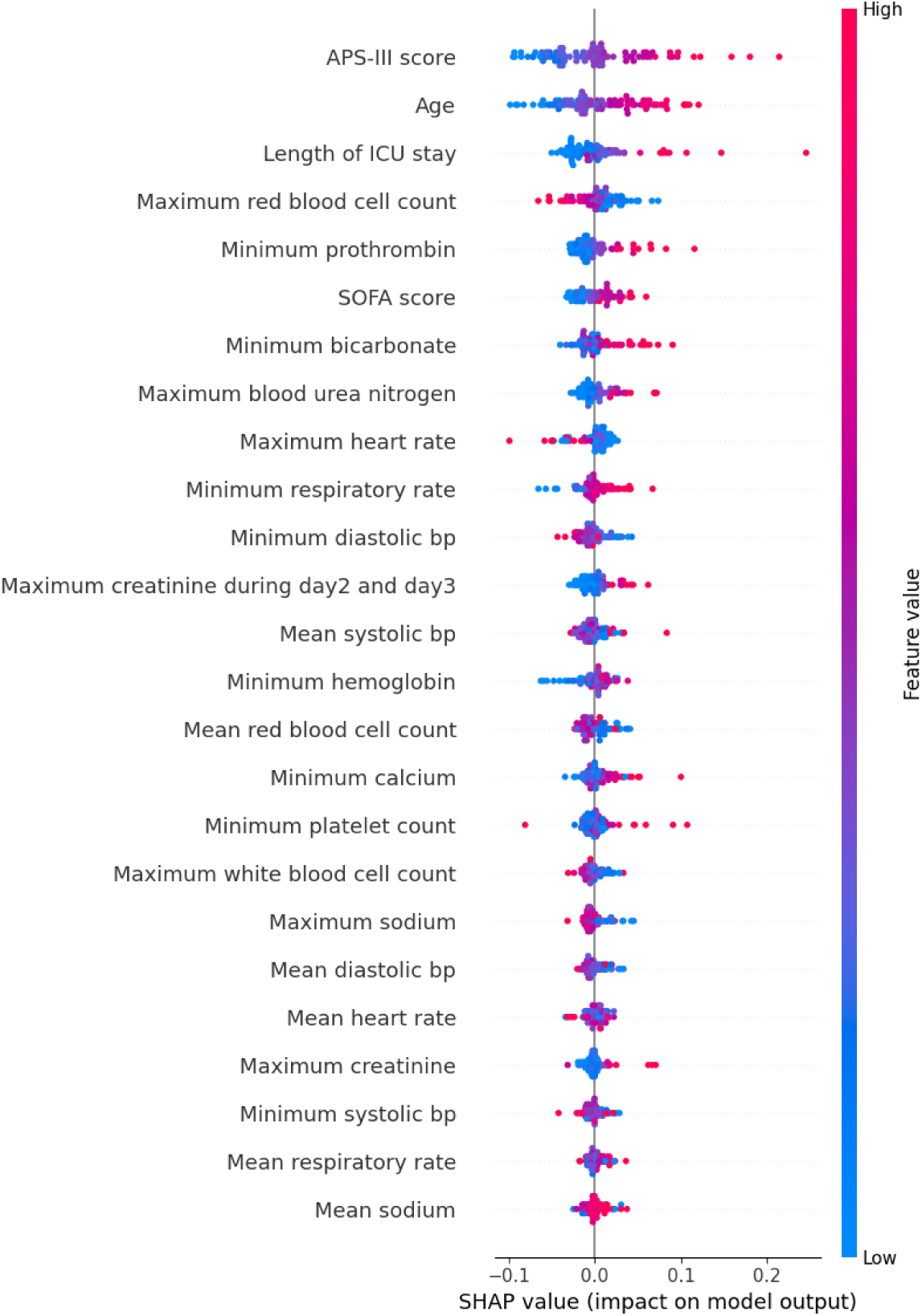
SHAP beeswarm plot of 1D CNN.

We further analyzed individual predictors with scatter plots. Figure 4(a) demonstrates that younger age (left side, negative SHAP values) contributes negatively to the model’s output, while older age (right side, positive SHAP values) contributes positively. Extreme ages appear to have a more significant impact on the model’s predictions, indicating that age has a monotonically increasing effect on the prediction of patient mortality. Figure 4(b) shows that SHAP values increase with the APS-III score, suggesting that a higher APS-III score correlates with an elevated probability of mortality, consistent with the score’s role in assessing patient risk.

**Figure 4.**
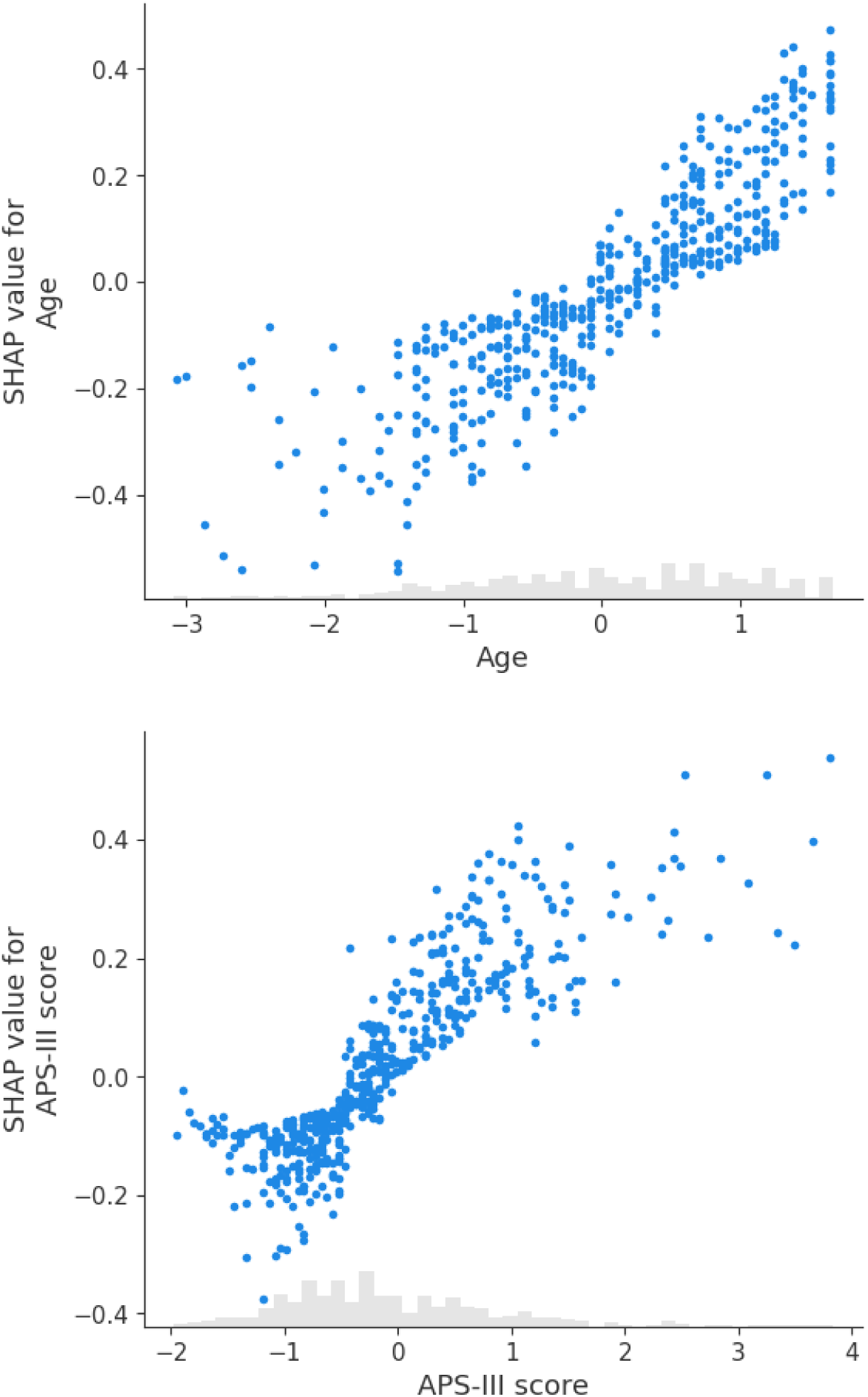
Scatter plots for SHAP values: (a) Age, and (b) APS-III score.

Finally, we used SHAP values to interpret two individual cases. Figure 5(a) displays the contribution of clinical measurements and patient characteristics to a positive mortality prediction, where age, APS-III score, and maximum blood urea nitrogen had the strongest positive contributions. Conversely, Figure 5(b) illustrates a case where the model predicted survival, with APS-III score, length of ICU stays, and SOFA score being the most significant factors in the negative prediction.

**Figure 5.**
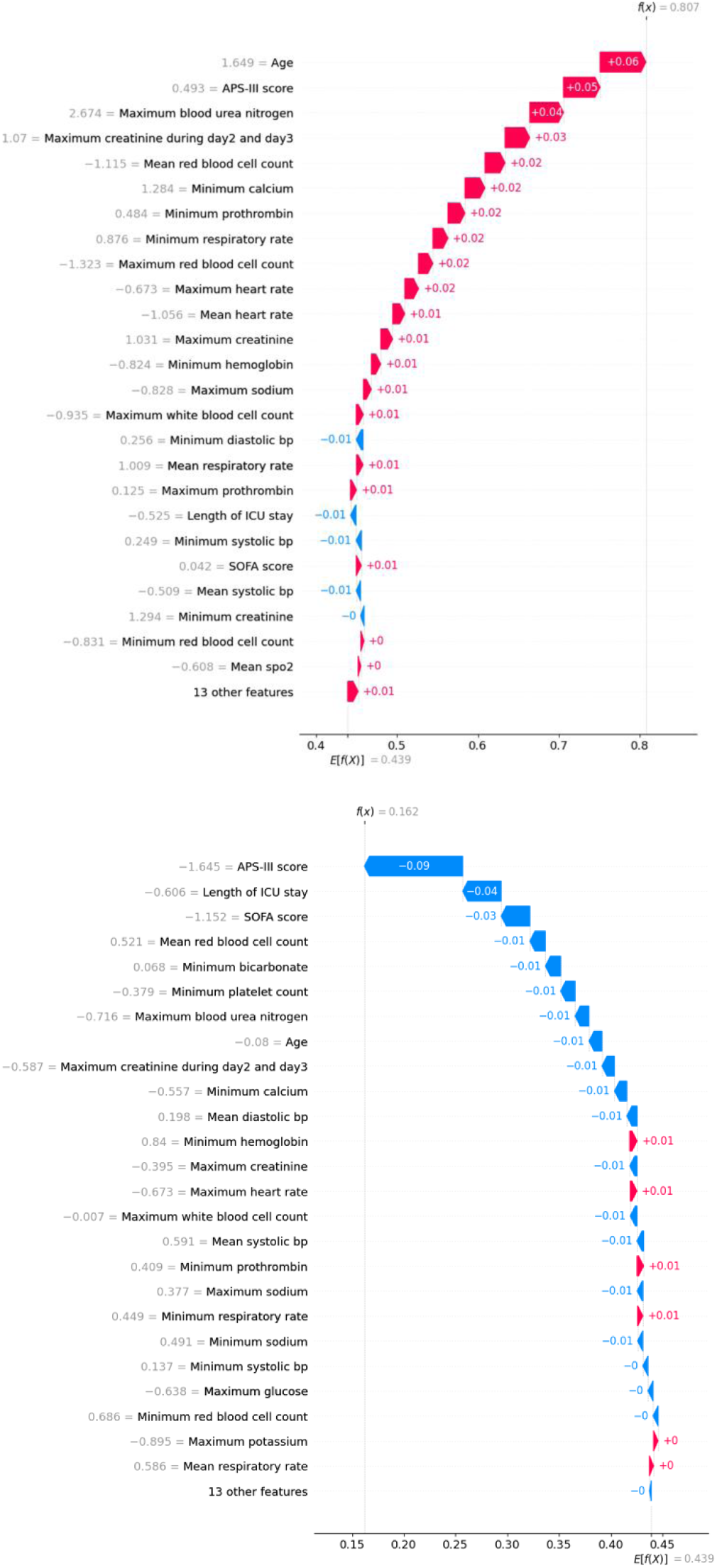
Waterfall plots for two individual cases: (a) True positive case, and (b) True negative case.

SHAP values were calculated for all models discussed in this paper.[26] Across models, we identified a consistent set of significant predictors for mortality in critically ill patients with hypertension. These predictors include minimum bicarbonate, APS-III score, maximum blood urea nitrogen, maximum creatinine during days 2 and 3, mean respiratory rate, minimum creatinine, maximum red blood cell count, length of ICU stay, age, minimum prothrombin, SOFA score, minimum systolic blood pressure, minimum diastolic blood pressure, and minimum hemoglobin.

## DISCUSSION

Accurate prognosis prediction is vital for patient-centered care, facilitating shared decision-making and informing treatment strategies. In this study, we evaluated various machine learning and deep learning models to predict mortality in critically ill patients with hypertension upon ICU admission. Our findings reveal that deep learning models, particularly the 1D CNN, consistently outperformed traditional machine learning models in predictive accuracy, as measured by AUC and other performance metrics. Specifically, the 1D CNN model, using an initial selection of predictors, achieved the highest AUC (0.7744), demonstrating its superior ability to distinguish between patients who survived and those who did not after ICU admission.

Our analysis also underscored the significance of specific clinical features, with the APS-III score, age, and length of ICU stay emerging as the most influential predictors of mortality. These results highlight the importance of comprehensive physiological assessments and detailed patient history in developing predictive models for critically ill populations.

The identification of key predictors such as APS-III score, age, and length of ICU stay carries significant implications for clinical practice.[27] These factors are routinely available in ICU settings, making the integration of the 1D CNN model into real-time clinical decision-making practical. With its high AUC and reliable positive predictive value, this model could assist healthcare providers in the early identification of high-risk patients, enabling more targeted interventions that may improve patient outcomes and optimize resource allocation.[28]

Moreover, the model’s ability to efficiently process large and complex datasets, coupled with its capacity for ongoing learning and adaptation, suggests that deep learning approaches could be instrumental in advancing precision medicine in critical care. This study demonstrates the effectiveness of both classic machine learning algorithms and modern deep learning models in predicting mortality among critically ill patients with hypertension. Our predictive models incorporated a diverse set of variables, leveraging professional medical knowledge by including SOFA and APS-III scores, which assess overall organ failure and the severity of illness. This approach enhanced the models’ ability to predict patient mortality more accurately.

Among the models evaluated, the 1D CNN exhibited the best performance, likely due to its architecture that combines the learning capabilities of neural networks with the efficiency of convolutional filters. The 1D CNN’s ability to capture local dependencies and recognize patterns within the data enabled it to outperform other models, including RNNs and LSTMs, which did not perform as well, possibly due to the lack of strong sequential dependencies in the predictors used.[29] Decision tree algorithms also performed well, consistent with existing literature, while support vector machines (SVMs) lagged behind, potentially due to the high-dimensional complexity of the dataset.

The 1D CNN prediction model has the potential to facilitate advanced clinical decision-making for critically ill patients with hypertension. The 1D CNN is designed to process sequential data, such as time series or any data with a temporal dimension. Its structure, which includes convolutional layers, activation functions, max pooling, and fully connected layers, is particularly suited for tasks requiring the recognition of local and positionally invariant patterns.[30] This makes 1D CNNs robust and efficient for analyzing large input sequences, eliminating the need for manual feature engineering, and ensuring robustness to shifts and variations in the input data. By reducing the number of parameters through shared weights and pooling, 1D CNNs are computationally efficient and can automatically learn to detect important features from raw data. This makes them particularly beneficial in complex clinical data scenarios, where data may not always be perfectly aligned or uniform. The model’s robustness to slight variations in input data is especially valuable in clinical contexts, where data consistency can vary.[30]

Our findings are consistent with previous research that has shown the utility of machine learning in predicting patient outcomes in the ICU.[31] However, this study advances the field by demonstrating the particular effectiveness of deep learning models, such as the 1D CNN, in handling the complex and multidimensional data typically found in ICU settings. Unlike traditional machine learning models, which often require extensive feature engineering, deep learning models can automatically discern intricate patterns and interactions within the data, leading to potentially more accurate predictions.[32]

The study also builds on prior work by using SHAP values to interpret the models, thereby providing transparency and insight into the decision-making process. Through the SHAP framework, we explored the impact of predictors across all employed models. This feature selection method has become widely recognized for explaining the effects of each feature on learning algorithms. Addressing one of the main criticisms of deep learning models—namely, their “black-box” nature—this approach elucidates the contributions of individual predictors to the model’s predictions. Such interpretability is crucial for gaining the trust of clinicians and ensuring that these models are used effectively and ethically in practice.

## LIMITATIONS

Despite its strengths, this study has several limitations. First, the study was conducted using a retrospective dataset, which may limit the generalizability of the findings to other patient populations or clinical settings. Future research should aim to validate these models prospectively in diverse ICU populations to confirm their utility and robustness in real-world settings. Second, although we employed SHAP values to enhance model interpretability, there remains a need for further research into the ethical implications of using such models in clinical practice. Issues such as algorithmic bias, patient consent, and the transparency of decision-making processes should be rigorously examined to ensure that these technologies are implemented responsibly.[33] Finally, while the models demonstrated good predictive performance, there is room for improvement. Exploring ensemble methods that combine multiple deep learning models or incorporating time-series data could further enhance predictive accuracy. Additionally, integrating multi-modal data sources, such as genomic information, clinical notes,[34] patient-generated health data,[35] and imaging data,[36] could provide a more comprehensive view of patient health, leading to even more personalized and precise predictions.[37]

## CONCLUSION

This study demonstrates the potential of deep learning models, particularly the 1D CNN, in predicting mortality in critically ill patients with hypertension in the ICU. The superior performance of these models, coupled with their ability to handle complex datasets and provide interpretable results, positions them as powerful tools for improving patient outcomes in critical care settings. However, ongoing research is needed to validate these findings in broader patient populations and to address the ethical challenges associated with the deployment of such technologies in clinical practice. By continuing to refine these models and ensuring their responsible use, we can move closer to realizing the promise of precision medicine in the ICU.

## CONTRIBUTION STATEMENT

ZZ contributed to data analyses and the writing of the manuscript. JY designed the study, contributed to the data analyses, and the writing of the manuscript. All authors read and approved the final version of the manuscript.

## DATA AVAILABILITY STATEMENT

All data are available at Medical Information Mart for Intensive Care: https://mimic.physionet.org/. The relevant code and analyses are available at: https://github.com/ZiyangZhang0511/predictingmortality/tree/main.

---

**Supplemental Table 1.** Bivariate association between outcome and predictors.

| Variable | odd ratio | p-value | 95% CI |
| --- | --- | --- | --- |
| Sex |  |  |  |
| Male (1) | 0.972 | 0.224 | [0.929, 1.018] |
| Female (0) | ref | ref | ref |
| Race |  |  |  |
| African American | ref | ref | ref |
| White | 0.745 | < 0.001 | [0.688, 0.806] |
| Hispanic | 0.552 | < 0.001 | [0.478, 0.639] |
| Other | 0.663 | < 0.001 | [0.578, 0.693] |
| Age, years | 1.029 | < 0.001 | [1.027, 1.031] |
| Minimum creatinine | 1.259 | < 0.001 | [1.236, 1.282] |
| Maximum creatinine | 1.201 | < 0.001 | [1.182, 1.219] |
| Maximum heart rate | 1.011 | < 0.001 | [1.010, 1.013] |
| Mean heart rate | 1.007 | < 0.001 | [1.006, 1.009] |
| Minimum systolic bp | 0.983 | < 0.001 | [0.982, 0.985] |
| Mean systolic bp | 0.990 | < 0.001 | [0.989, 0.992] |
| Minimum diastolic bp | 0.968 | < 0.001 | [0.966, 0.970] |
| Mean diastolic bp | 0.980 | < 0.001 | [0.978, 0.982] |
| Minimum SpO2 | 0.996 | 0.222 | [0.989, 1.003] |
| Mean SpO2 | 0.993 | 0.263 | [0.980, 1.006] |
| Minimum hemoglobin | 0.894 | < 0.001 | [0.885, 0.903] |
| Minimum prothrombin | 1.062 | < 0.001 | [1.057, 1.066] |
| Maximum prothrombin | 1.030 | < 0.001 | [1.027, 1.033] |
| Minimum respiratory rate | 1.036 | < 0.001 | [1.031, 1.042] |
| Maximum respiratory rate | 1.016 | < 0.001 | [1.013, 1.019] |
| Mean respiratory rate | 1.062 | < 0.001 | [1.056, 1.069] |
| Maximum glucose | 1.001 |  | [1.001, 1.001] |
| Minimum platelet count | 1.000 | 0.382 | [1.000, 1.000] |
| Minimum calcium | 0.959 | 0.003 | [0.932, 0.986] |
| Minimum bicarbonate | 1.009 | 0.001 | [1.004, 1.015] |
| Maximum potassium | 1.122 | < 0.001 | [1.089, 1.156] |
| Maximum blood urea nitrogen | 1.025 | < 0.001 | [1.023, 1.026] |
| Maximum red blood cell count | 0.594 | < 0.001 | [0.574, 0.615] |
| Minimum red blood cell count | 0.730 | < 0.001 | [0.708, 0.753] |
| Mean red blood cell count | 0.640 | < 0.001 | [0.619, 0.662] |
| Maximum white blood cell count | 1.003 | 0.002 | [1.001, 1.006] |
| Maximum sodium | 1.000 | 0.964 | [0.996, 1.004] |
| Minimum sodium | 0.995 | < 0.001 | [0.994, 0.995] |
| Mean sodium | 0.988 | < 0.001 | [0.985, 0.990] |
| Length of ICU stay (days) | 1.136 | < 0.001 | [1.128, 1.144] |
| SOFA score | 1.037 | < 0.001 | [1.035, 1.038] |
| APS-III score | 1.034 | < 0.001 | [1.030, 1.038] |

**Supplemental Table 2.** Multivariable logistic regression of backward selection.

| **Variables** | **Adjusted Odds ratio** | **P-value** | **95% confidence interval** |
| --- | --- | --- | --- |
| Sex |  |  |  |
| Male (1) | 0.0586 | 0.028 | [0.006, 0.111] |
| Female (0) | ref | ref | ref |
| Race |  |  |  |
| African American | ref | ref | ref |
| White | -0.3904 | < 0.001 | [-0.482, -0.299] |
| Hispanic | -0.4724 | < 0.001 | [-0.634, -0.311] |
| Other | -0.4935 | < 0.001 | [-0.597, -0.390] |
| Ag, years | 1.5370 | < 0.001 | [1.494, 1.582] |
| Minimum creatinine | 1.1579 | 0.006 | [1.043, 1.286] |
| Maximum creatinine | 0.7360 | < 0.001 | [0.662, 0.818] |
| Mean heart rate | 1.0414 | 0.005 | [1.012, 1.072] |
| Minimum systolic bp | 0.8688 | < 0.001 | [0.836, 0.904] |
| Mean systolic bp | 1.0605 | 0.004 | [1.019, 1.104] |
| Minimum hemoglobin | 0.8545 | < 0.001 | [0.807, 0.905] |
| Minimum prothrombin | 1.1912 | < 0.001 | [1.156, 1.227] |
| Minimum respiratory rate | 1.0771 | < 0.001 | [1.043, 1.113] |
| Maximum respiratory rate | 0.9295 | < 0.001 | [0.898, 0.962] |
| Mean respiratory rate | 1.0900 | < 0.001 | [1.046, 1.135] |
| Maximum glucose | 1.0479 | 0.001 | [1.019, 1.077] |
| Minimum calcium | 1.0327 | 0.027 | [1.004, 1.063] |
| Minimum bicarbonate | 1.1297 | < 0.001 | [1.098, 1.162] |
| Maximum blood urea nitrogen | 1.2297 | < 0.001 | [1.184, 1.277] |
| Maximum red blood cell count | 0.5673 | < 0.001 | [0.534, 0.601] |
| Minimum red blood cell count | 1.7058 | < 0.001 | [1.571, 1.852] |
| SOFA Score | 1.0711 | < 0.001 | [1.032, 1.112] |
| APS-III Score | 1.6294 | < 0.001 | [1.565, 1.696] |
| Length of ICU stay | 1.1565 | < 0.001 | [1.126, 1.188] |

**Supplemental Table 3.**
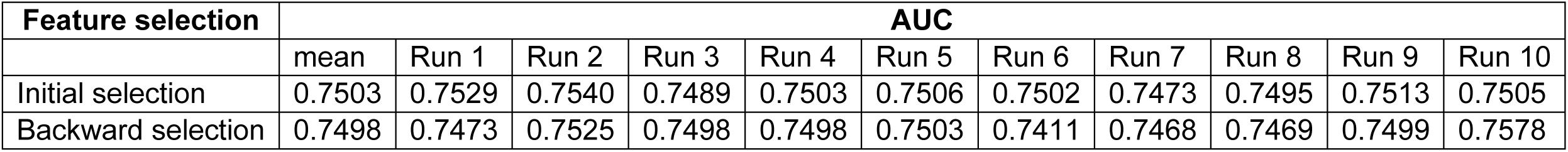
Multivariable logistic regression AUC.

